# Patient- and Ward-Level Determinants of Psychosomatic-Psychiatric Consultations for Mentally Distressed Inpatients from Medical Hospitals: Findings from the SomPsyNet Stepped-Wedge-Trial

**DOI:** 10.64898/2026.02.13.26346221

**Authors:** Gunther Meinlschmidt, Alexander Frick, Iris Baenteli, Christina Karpf, Anja Studer, Sanaa Bahmane, Nikolia Cicic, David Buechel, Lukas Ebner, Marco Bachmann, Andreas Dörner, Sibil Tschudin, Sarah Trost, Kaspar Wyss, Günther Fink, Matthias Schwenkglenks, Rainer Schaefert, the SomPsyNet consortium

## Abstract

**Background:** Up to one-third of medical inpatients experience clinically relevant mental distress, yet many remain untreated. Stepped and collaborative care (SCC) models may improve access to mental health care, but predictors of service uptake are unclear. We examined patient- and ward-level predictors of psychosomatic-psychiatric consultation (PPC).

**Methods:** We analyzed data from SomPsyNet, a stepped-wedge cluster randomized trial targeting SOMatic inpatients across three Swiss tertiary hospitals, to prevent PSYchosocial distress by a care NETwork. Analyses focused on inpatients screening positive for mental distress. Multiple-imputed logistic regressions assessed predictors of four sequential service-use stages: PPC considered, offered, accepted, and received.

**Results:** Among 589 distressed patients, 93.9% were offered PPC, 63.1% accepted, and 83.9% of acceptors received PPC, yielding a 50% overall receipt rate. Patients without Swiss citizenship showed higher odds of acceptance (odds ratio [OR]=1.82 [1.10, 3.00]) and eventual receipt (OR=1.62 [1.01, 2.62]). Being in a Geriatric ward facilitated PCC uptake, while patients from gynecology showed reduced progression through the care pathway. Age, gender, income, education, marital status, and living arrangement showed no statistically robust associations.

**Conclusions:** Almost two-thirds of mentally distressed medical hospital inpatients accepted an offered PPC, indicating high acceptability. About half ultimately received a consultation, highlighting substantial attrition along the SCC pathway. Ward specialty and nationality were key determinants of PPC uptake. These findings suggest that proactive, ward-oriented consultation-liaison models embedded in routine inpatient care may improve timely and equitable access to mental healthcare, including for migrant and minority patients who are otherwise less likely to access such care.

**Highlights:**

- Psychosomatic-psychiatric consultation pathway of medically hospitalized inpatients
- 63% accepted such a consultation when offered; overall 50% reached receipt.
- Non-Swiss nationality increased odds of acceptance (OR 1.8) and receipt (OR 1.6).
- Patients at geriatrics wards showed higher, at gynecology wards lower transitions.
- Results support low-threshold, ward-oriented consultation-liaison models.

## 1. Introduction

Medically hospitalized inpatients commonly experience considerable mental or psychosocial distress, with approximately 30% developing clinically relevant symptoms of depression, anxiety, or somatic symptom distress during their hospital stay [2,3].

Such distress, if unrecognized or undertreated, may amplify morbidity, increase (re-) hospitalization rates, prolonged hospital stays, and elevate healthcare costs [4,5]. To address these challenges, stepped and collaborative care models (SCCMs) have increasingly been adopted in multiple medical fields, yet in most cases in primary care and ambulatory or outpatient settings [6]. Key attributes – systematic distress identification, structured care pathways, and close coordination between somatic and mental health services – are endorsed in guidelines (e.g., American Society of Clinical Oncology [7]; hospital distress-screening task forces [8], and supported by meta-analyses [9]. Notably, most SCCMs also involve a dedicated care coordinator based in primary care, whereas few models have the coordinator embedded within a medical hospital inpatient setting.

The SomPsyNet project aims at patients from SOMatic hospitals and promotes the prevention of PSYchosocial distress by establishing a structured care NETwork. Therefore, it was designed to improve the detection and management of psychosocial distress among medical hospital inpatients through an SCCM in selected medical hospitals in Switzerland [10,11]. The SomPsyNet initiative, implemented in Basel-Stadt, combines early screening for depression, anxiety, somatic distress, and other psychosocial stressors with stepped psychosomatic-psychiatric interventions tailored to patients’ needs [12]. Beyond improving patients’ health-related quality of life, these approaches may potentially reduce detrimental outcomes such as extended morbidity and recurrent hospital admissions [4]. The stepped-wedge cluster randomized design of SomPsyNet allowed a sequential roll-out of the model into routine care – transitioning clusters within wards from treatment-as-usual (no screening) to an intervention phase with psychosocial screening and subsequent psychosomatic-psychiatric consultations (PCC) [12].

Although stepped and collaborative care frameworks show promise, it remains unclear what proportion of distressed medically hospitalized inpatients ultimately receive PCCs, and which patient- or ward-level factors influence progression through the successive stages of this care pathway. Only few studies have investigated how many patients actually accept a PCC, and which specific patient or institutional characteristics predict whether a potentially distressed patient will proceed through the different stages of care [13]. Limited evidence suggests that institutional factors – such as ward culture or staff attitudes – may affect psychosocial service uptake in medical settings [11]. Overall, findings have been mixed, and many healthcare systems still struggle to ensure consistent referral and uptake of mental health interventions among medically ill patients.

Against this backdrop, the SomPsyNet project offers a valuable opportunity to explore individual- and hospital-level influences on the actual receipt of psychosomatic-psychiatric care within a large-scale SCCM. The present study aimed to i) quantify the proportion of medically hospitalized inpatients transitioning through each key stage of psychosomatic-psychiatric care – from mere *consideration* of a consultation to *offering*, *accepting*, and ultimately *receiving* it; and ii) identify sociodemographic and ward-level predictors (e.g., age, nationality, medical ward specialty) that increase or decrease the likelihood of patients transitioning between stages in the care pathway.

By examining these transitions in a real-world setting, the study contributes to the broader literature on how stepped and collaborative care can be operationalized to address psychosocial distress in medical hospital inpatient settings. The resulting insights may help refine SCCMs and inform targeted interventions to optimize mental health service uptake in this population.

## 2. Methods

### 2.1. Study setting and stepped-wedge cluster randomized design

We focus on data from a subsample of the SomPsyNet study, conducted in three Swiss tertiary medical hospitals [12], of which a baseline description of the study population has been submitted elsewhere [3]. The SomPsyNet project is managed by the Department of Psychosomatic Medicine at the University Hospital Basel (UHB) and the Medical Services of the Department of Health Basel-Stadt (GD), in collaboration with Bethesda Hospital (BESP), the c, and other healthcare partners. The study aimed to investigate SCCM for medically hospitalized inpatients. Detailed information on the larger SomPsyNet project’s setting and design is available elsewhere [12]. The stepped-wedge cluster randomized trial was conducted in three phases: i) standard treatment without distress screening; ii) treatment as usual plus distress screening but no intervention; and iii) full SCCM with recommended intervention for patients identified with mental distress. We restricted our analyses to inpatients who screened positive for distress during the final phase iii), defined as having at least one indicator of mental distress above the relevant cutoff (e.g., clinically significant depression, anxiety, or somatic distress); for details, see the published study protocol [12], and baseline analysis of the SomPsyNet study data [3].

### 2.2. Study registration and ethics

The SomPsyNet study adheres to the Declaration of Helsinki, the Human Research Act (HRA), and local regulations. Participation was voluntary, with written informed consent required from all participants. Ethical approval was granted by the *Ethikkommission Nordwest- und Zentralschweiz* (EKNZ; No. 2019–01724), including four subsequent amendments reflecting updates to the study. The study has been registered with the Swiss National Clinical Trials Portal (SNCTP) and ClinicalTrials.gov (NCT04269005).

### 2.3. Inclusion and exclusion criteria

Recruitment of SomPsyNet participants included in the here reported analyses was carried out between 09-06-2020 to 16-12-2022, across the UHB, BESP, and UAFP.

Patients admitted to the participating wards were systematically identified for enrolment via the hospitals’ electronic management systems and direct coordination with ward personnel. Each potential participant’s medical records were reviewed, and discussions were held with patients to confirm eligibility. All new admissions were considered unless they met exclusion criteria. Eligible patients were informed about the study and if they gave their consent, their data were included. Only the first admission per patient was analyzed. If a participant withdrew, data collection ceased, and previously collected data were excluded. Eligible participants were those hospitalized in the participating wards during the study recruitment period. Study inclusion criterion was hospitalization in a ward actively participating in SomPsyNet.

Exclusion criteria included being under 18 years of age, inability to understand or speak German, incapacity to consent, severe medical conditions precluding participation, risk of suicidality, a condition with already implemented specific psychosocial support (e.g., oncological conditions, hospitalization for gender-affirming interventions), prior SomPsyNet participation, active COVID-19 infection, or medical supervision from a non-participating ward despite physical location in a participating ward. Although multiple languages were initially considered, data collection proceeded solely in German. Patients’ pathways to inclusion varied, encompassing direct admissions from home or transfers from other hospitals, the emergency department, intensive care units, or other wards.

### 2.4. Study variables

The full set of data collected has been described elsewhere [12]. For the present analyses, we collected a comprehensive set of sociodemographic characteristics at baseline, including age, gender, nationality, household income, marital status, living arrangement, highest education level, and employment situation. The medical ward specialty was defined by the key medical specialty of the ward at which the patients were hospitalized. These variables were captured through standardized questionnaires and, when required, hospital records.

As events to predict, we focused on four service-uptake transitions across the different stages towards providing PPC for those distressed:

1. Stage 0 to stage 1: From “PPC considered” (the patient was identified as needing a PPC based on a positive distress screening) to “PPC offered.”
2. Stage 1 to stage 2: From “PPC offered” to “PPC accepted.”
3. Stage 2 to stage 3: From “PPC accepted” to “PPC received.”
4. Stage 0 to stage 3: From “PPC considered” to “PPC received.” (an overall transition summarizing progression through all stages)

All individuals who met the inclusion criteria at each starting stage of the transition were included in the corresponding analyses.

### 2.5. Data collection

After obtaining informed consent, patients completed a baseline questionnaire, primarily via a tablet-based software system (‘heartbeat one’, Heartbeat Medical Solutions GmbH, Berlin, Germany). Paper-pencil and staff-assisted formats were available when necessary, and any paper-pencil data were entered into ‘heartbeat one’ by study staff. All data were then transferred to the secuTrial® database for verification and resolution of discrepancies. Sociodemographic variables and medical ward specialties were classified according to Table 1.

**Table 1.** Sociodemographic characteristics of the study sample (PPC considered, *N* = 589)

|  | Overall |  |  |
| --- | --- | --- | --- |
|  | N | % | [CI LB; CI UB] |
| **Age (in years)** |  |  |  |
| up to 30 | 50 | 8.49 | [3.13; 21.04] |
| 30 - 39 | 72 | 12.22 | [5.71; 24.27] |
| 40 - 49 | 70 | 11.88 | [6.77; 20.04] |
| 50 - 59 | 92 | 15.62 | [10.06; 23.45] |
| 60 - 69 | 92 | 15.62 | [9.36; 24.91] |
| 70 - 79 | 125 | 21.22 | [13.82; 31.15] |
| 80 and more | 88 | 14.94 | [6.61; 30.36] |
| **Gender** |  |  |  |
| Female | 316 | 53.65 | [34.17; 72.08] |
| Male or other | 273 | 46.35 | [27.92; 65.83] |
| **Household income (in CHF per month)<sup>a</sup>** |  |  |  |
| less than 4,500 | 146 | 35.1 | [29.32; 41.35] |
| 4,500 - 5,999 | 118 | 28.37 | [25.32; 31.62] |
| 6,000 - 8,999 | 84 | 20.19 | [16.14; 24.95] |
| 9,000 or more | 68 | 16.35 | [13.60; 19.52] |
| **Nationality** |  |  |  |
| Swiss citizenship | 467 | 79.29 | [72.03; 85.05] |
| No Swiss citizenship | 122 | 20.71 | [14.95; 27.97] |
| **Marital status<sup>b</sup>** |  |  |  |
| Divorced or dissolved partnership / widowed | 153 | 26.24 | [15.96; 39.99] |
| Married or partnered | 249 | 42.71 | [34.29; 51.57] |
| Single or unmarried | 181 | 31.05 | [19.39; 45.74] |
| **Living arrangement<sup>c</sup>** |  |  |  |
| With spouse/partner/family/friends | 332 | 66.27 | [60.48; 71.61] |
| Alone / in a nursing home / hospital or other long-term care facility / other | 169 | 33.73 | [28.39; 39.52] |
| **Highest education level<sup>d</sup>** |  |  |  |
| Did not attend school / primary level / not applicable / lower secondary level I / other | 90 | 15.41 | [11.74; 19.96] |
| Upper secondary level II | 268 | 45.89 | [39.57; 52.35] |
| Tertiary level | 226 | 38.7 | [31.01; 46.99] |
| **Employment<sup>e</sup>** |  |  |  |
| Employed or in training | 179 | 39.43 | [25.57; 55.22] |
| Pension recipient | 184 | 40.53 | [25.55; 57.50] |
| Disability, accident or physical illness | 52 | 11.45 | [9.07; 14.37] |
| Homemaker/ unemployed or looking for a job/ in military service / other | 39 | 8.59 | [5.92; 12.30] |
| **Medical specialty** |  |  |  |
| Geriatrics | 60 | 10.19 | [0.82; 60.90] |
| Gynecology | 105 | 17.83 | [2.39; 65.75] |
| Internal medicine | 318 | 53.99 | [17.18; 86.91] |
| Rehabilitation | 67 | 11.38 | [0.93; 63.82] |
| Rheumatology | 39 | 6.62 | [0.52; 49.22] |

<sup>a</sup>173 missing values; <sup>b</sup>6 missing values; <sup>c</sup>88 missing values; <sup>d</sup>5 missing values; <sup>e</sup>135 missing values.
Abbreviations: CHF, Swiss Francs; CI, 95% confidence interval; LB, lower bound; PPC, psychosomatic-psychiatric consultation; UB, upper bound.
Note. Categories with n < 10 are not shown for data protection reasons.

### 2.6. Patient and public involvement

SomPsyNet established a patient participation committee with patient representatives involved from the project’s inception, including grant preparation and study design. Their input improved study materials, informed consent processes, enrolment strategies, and coordination issues of follow-up care, and they contributed to disseminating findings.

### 2.7. Statistical methods

We began by generating frequency counts and percentages for key sociodemographic variables and medical ward specialty. We estimated 95% Confidence Intervals (CIs) for proportions with continuity correction. Then we produced descriptive summaries in the form of cross-tabulations of demographic characteristics against the relative frequency of each service-use stage transition. We applied robust standard errors clustered by ward/ hospital unit within the survey design, accounting for potential intracluster correlation and variance inflation.

Before analysis, we addressed missing values in the predictor set. Little’s χ² test showed the data were not Missing Completely at Random (MCAR; *p*<0.05). To explore whether a Missing at Random (MAR) mechanism was plausible, we created binary indicators of missingness and regressed them on the observed covariates; significant predictors of missingness supported plausibility of MAR. We therefore carried out Multiple Imputation by Chained Equations (MICE) with Stata’s *mi* framework, generating 100 imputations to minimize Monte-Carlo error. We used multinomial logistic regression for nominal variables and ordinal logistic regression for ordinal variables.

We conducted separate logistic regression analyses to examine predictors of successfully transitioning across each of the service use stages, and of all service use stages combined (see above). Hence, we estimated four separate models. As predictors, we included the variables listed in Table 2. Thereby, each predictor’s association with the outcome was adjusted for the other predictors in the model. Further, we performed tests for trends for the ordinal data, age, household income, and highest educational level to identify linear patterns or significant trends within these variables with ordered categories.

**Table 2.**
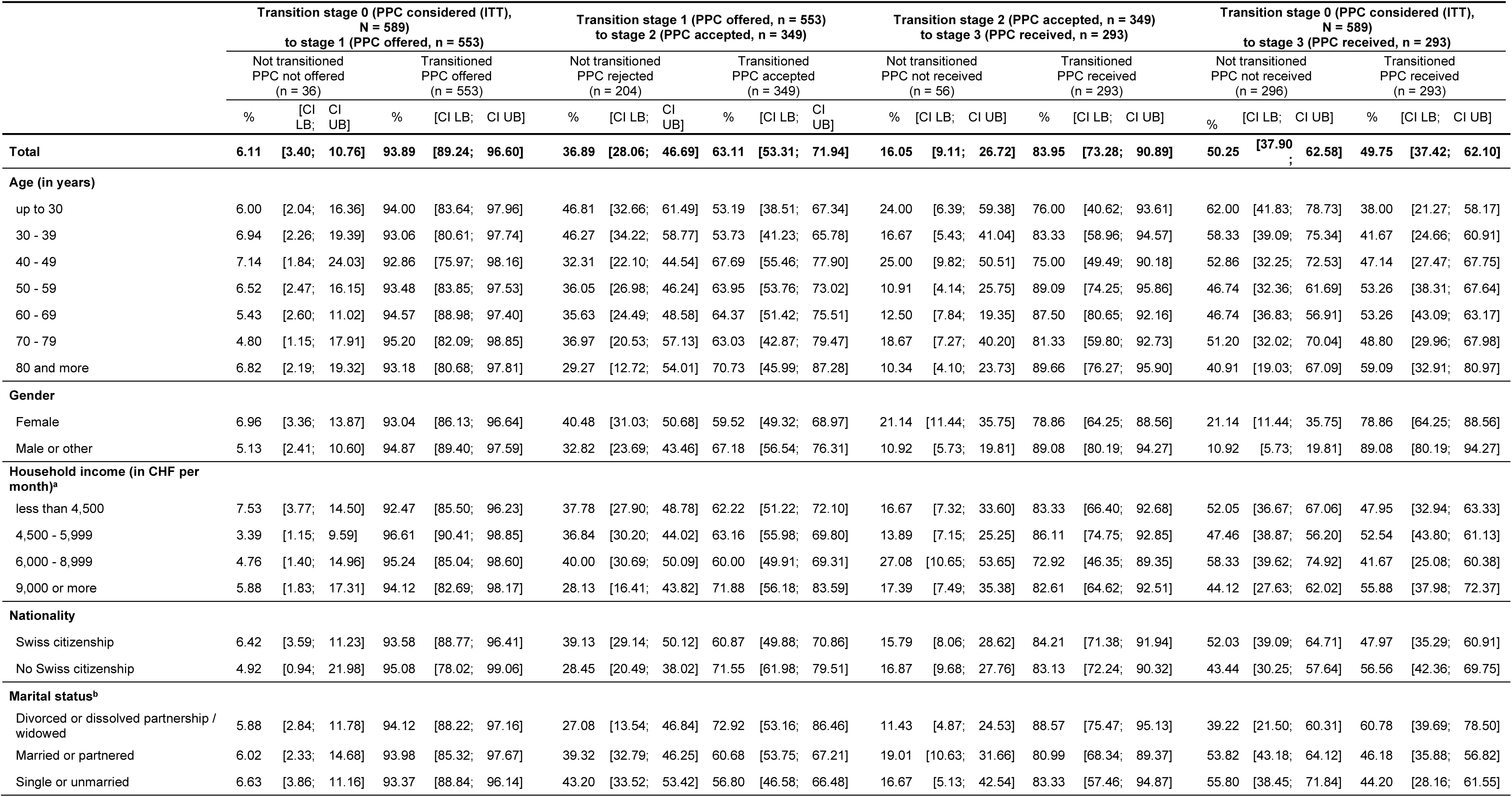
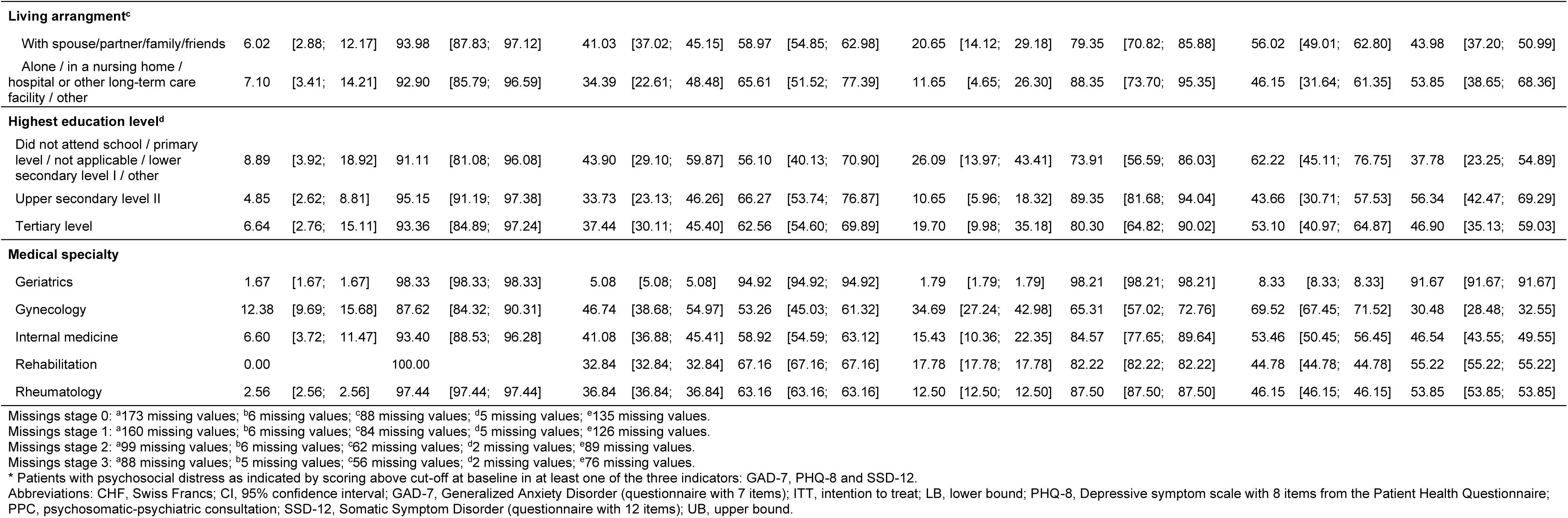
Frequency of transitioning across the different PPC stages stratified by sociodemographic characteristics.

|  | Transition stage 0 (PPC considered (ITT),<br>N = 589)<br>to stage 1 (PPC offered, n = 553) |  |  |  |  |  | Transition stage 1 (PPC offered, n = 553)<br>to stage 2 (PPC accepted, n = 349) |  |  |  |  |  | Transition stage 2 (PPC accepted, n = 349)<br>to stage 3 (PPC received, n = 293) |  |  |  |  |  | Transition stage 0 (PPC considered (ITT),<br>N = 589)<br>to stage 3 (PPC received, n = 293) |  |  |  |  |
| --- | --- | --- | --- | --- | --- | --- | --- | --- | --- | --- | --- | --- | --- | --- | --- | --- | --- | --- | --- | --- | --- | --- | --- |
|  | Not transitioned<br>PPC not offered<br>(n = 36) |  |  | Transitioned<br>PPC offered<br>(n = 553) |  |  | Not transitioned<br>PPC rejected<br>(n = 204) |  |  | Transitioned<br>PPC accepted<br>(n = 349) |  |  | Not transitioned<br>PPC not received<br>(n = 56) |  |  | Transitioned<br>PPC received<br>(n = 293) |  |  | Not transitioned<br>PPC not received<br>(n = 296) |  |  | Transitioned<br>PPC received<br>(n = 293) |  |
|  | % | [CI<br>LB; CI<br>UB] |  | % | [CI LB; CI UB] |  | % | [CI LB; CI UB] |  | % | [CI LB; CI UB] |  | % | [CI LB; CI UB] |  | % | [CI LB; CI UB] |  | % | [CI LB; CI UB] |  | % | [CI LB; CI UB] |
| **Total** | **6.11** | **[3.40; 10.76]** |  | **93.89** | **[89.24; 96.60]** |  | **36.89** | **[28.06; 46.69]** |  | **63.11** | **[53.31; 71.94]** |  | **16.05** | **[9.11; 26.72]** |  | **83.95** | **[73.28; 90.89]** |  | **50.25** | **[37.90; 62.58]** |  | **49.75** | **[37.42; 62.10]** |
| **Age (in years)** |  |  |  |  |  |  |  |  |  |  |  |  |  |  |  |  |  |  |  |  |  |  |  |
| up to 30 | 6.00 | [2.04; 16.36] |  | 94.00 | [83.64; 97.96] |  | 46.81 | [32.66; 61.49] |  | 53.19 | [38.51; 67.34] |  | 24.00 | [6.39; 59.38] |  | 76.00 | [40.62; 93.61] |  | 62.00 | [41.83; 78.73] |  | 38.00 | [21.27; 58.17] |
| 30 - 39 | 6.94 | [2.26; 19.39] |  | 93.06 | [80.61; 97.74] |  | 46.27 | [34.22; 58.77] |  | 53.73 | [41.23; 65.78] |  | 16.67 | [5.43; 41.04] |  | 83.33 | [58.96; 94.57] |  | 58.33 | [39.09; 75.34] |  | 41.67 | [24.66; 60.91] |
| 40 - 49 | 7.14 | [1.84; 24.03] |  | 92.86 | [75.97; 98.16] |  | 32.31 | [22.10; 44.54] |  | 67.69 | [55.46; 77.90] |  | 25.00 | [9.82; 50.51] |  | 75.00 | [49.49; 90.18] |  | 52.86 | [32.25; 72.53] |  | 47.14 | [27.47; 67.75] |
| 50 - 59 | 6.52 | [2.47; 16.15] |  | 93.48 | [83.85; 97.53] |  | 36.05 | [26.98; 46.24] |  | 63.95 | [53.76; 73.02] |  | 10.91 | [4.14; 25.75] |  | 89.09 | [74.25; 95.86] |  | 46.74 | [32.36; 61.69] |  | 53.26 | [38.31; 67.64] |
| 60 - 69 | 5.43 | [2.60; 11.02] |  | 94.57 | [88.98; 97.40] |  | 35.63 | [24.49; 48.58] |  | 64.37 | [51.42; 75.51] |  | 12.50 | [7.84; 19.35] |  | 87.50 | [80.65; 92.16] |  | 46.74 | [36.83; 56.91] |  | 53.26 | [43.09; 63.17] |
| 70 - 79 | 4.80 | [1.15; 17.91] |  | 95.20 | [82.09; 98.85] |  | 36.97 | [20.53; 57.13] |  | 63.03 | [42.87; 79.47] |  | 18.67 | [7.27; 40.20] |  | 81.33 | [59.80; 92.73] |  | 51.20 | [32.02; 70.04] |  | 48.80 | [29.96; 67.98] |
| 80 and more | 6.82 | [2.19; 19.32] |  | 93.18 | [80.68; 97.81] |  | 29.27 | [12.72; 54.01] |  | 70.73 | [45.99; 87.28] |  | 10.34 | [4.10; 23.73] |  | 89.66 | [76.27; 95.90] |  | 40.91 | [19.03; 67.09] |  | 59.09 | [32.91; 80.97] |
| **Gender** |  |  |  |  |  |  |  |  |  |  |  |  |  |  |  |  |  |  |  |  |  |  |  |
| Female | 6.96 | [3.36; 13.87] |  | 93.04 | [86.13; 96.64] |  | 40.48 | [31.03; 50.68] |  | 59.52 | [49.32; 68.97] |  | 21.14 | [11.44; 35.75] |  | 78.86 | [64.25; 88.56] |  | 21.14 | [11.44; 35.75] |  | 78.86 | [64.25; 88.56] |
| Male or other | 5.13 | [2.41; 10.60] |  | 94.87 | [89.40; 97.59] |  | 32.82 | [23.69; 43.46] |  | 67.18 | [56.54; 76.31] |  | 10.92 | [5.73; 19.81] |  | 89.08 | [80.19; 94.27] |  | 10.92 | [5.73; 19.81] |  | 89.08 | [80.19; 94.27] |
| **Household income (in CHF per month)<sup>a</sup>** |  |  |  |  |  |  |  |  |  |  |  |  |  |  |  |  |  |  |  |  |  |  |  |
| less than 4,500 | 7.53 | [3.77; 14.50] |  | 92.47 | [85.50; 96.23] |  | 37.78 | [27.90; 48.78] |  | 62.22 | [51.22; 72.10] |  | 16.67 | [7.32; 33.60] |  | 83.33 | [66.40; 92.68] |  | 52.05 | [36.67; 67.06] |  | 47.95 | [32.94; 63.33] |
| 4,500 - 5,999 | 3.39 | [1.15; 9.59] |  | 96.61 | [90.41; 98.85] |  | 36.84 | [30.20; 44.02] |  | 63.16 | [55.98; 69.80] |  | 13.89 | [7.15; 25.25] |  | 86.11 | [74.75; 92.85] |  | 47.46 | [38.87; 56.20] |  | 52.54 | [43.80; 61.13] |
| 6,000 - 8,999 | 4.76 | [1.40; 14.96] |  | 95.24 | [85.04; 98.60] |  | 40.00 | [30.69; 50.09] |  | 60.00 | [49.91; 69.31] |  | 27.08 | [10.65; 53.65] |  | 72.92 | [46.35; 89.35] |  | 58.33 | [39.62; 74.92] |  | 41.67 | [25.08; 60.38] |
| 9,000 or more | 5.88 | [1.83; 17.31] |  | 94.12 | [82.69; 98.17] |  | 28.13 | [16.41; 43.82] |  | 71.88 | [56.18; 83.59] |  | 17.39 | [7.49; 35.38] |  | 82.61 | [64.62; 92.51] |  | 44.12 | [27.63; 62.02] |  | 55.88 | [37.98; 72.37] |
| **Nationality** |  |  |  |  |  |  |  |  |  |  |  |  |  |  |  |  |  |  |  |  |  |  |  |
| Swiss citizenship | 6.42 | [3.59; 11.23] |  | 93.58 | [88.77; 96.41] |  | 39.13 | [29.14; 50.12] |  | 60.87 | [49.88; 70.86] |  | 15.79 | [8.06; 28.62] |  | 84.21 | [71.38; 91.94] |  | 52.03 | [39.09; 64.71] |  | 47.97 | [35.29; 60.91] |
| No Swiss citizenship | 4.92 | [0.94; 21.98] |  | 95.08 | [78.02; 99.06] |  | 28.45 | [20.49; 38.02] |  | 71.55 | [61.98; 79.51] |  | 16.87 | [9.68; 27.76] |  | 83.13 | [72.24; 90.32] |  | 43.44 | [30.25; 57.64] |  | 56.56 | [42.36; 69.75] |
| **Marital status<sup>b</sup>** |  |  |  |  |  |  |  |  |  |  |  |  |  |  |  |  |  |  |  |  |  |  |  |
| Divorced or dissolved partnership / widowed | 5.88 | [2.84; 11.78] |  | 94.12 | [88.22; 97.16] |  | 27.08 | [13.54; 46.84] |  | 72.92 | [53.16; 86.46] |  | 11.43 | [4.87; 24.53] |  | 88.57 | [75.47; 95.13] |  | 39.22 | [21.50; 60.31] |  | 60.78 | [39.69; 78.50] |
| Married or partnered | 6.02 | [2.33; 14.68] |  | 93.98 | [85.32; 97.67] |  | 39.32 | [32.79; 46.25] |  | 60.68 | [53.75; 67.21] |  | 19.01 | [10.63; 31.66] |  | 80.99 | [68.34; 89.37] |  | 53.82 | [43.18; 64.12] |  | 46.18 | [35.88; 56.82] |
| Single or unmarried | 6.63 | [3.86; 11.16] |  | 93.37 | [88.84; 96.14] |  | 43.20 | [33.52; 53.42] |  | 56.80 | [46.58; 66.48] |  | 16.67 | [5.13; 42.54] |  | 83.33 | [57.46; 94.87] |  | 55.80 | [38.45; 71.84] |  | 44.20 | [28.16; 61.55] |

|  |  |  |  |  |  |  |  |  |  |  |  |  |  |  |  |  |  |  |  |  |  |  |  |  |
| --- | --- | --- | --- | --- | --- | --- | --- | --- | --- | --- | --- | --- | --- | --- | --- | --- | --- | --- | --- | --- | --- | --- | --- | --- |
| Living arrangement<sup>c</sup> |  |  |  |  |  |  |  |  |  |  |  |  |  |  |  |  |  |  |  |  |  |  |  |  |
| With spouse/partner/family/friends | 6.02 | [2.88; | 12.17] | 93.98 | [87.83; | 97.12] | 41.03 | [37.02; | 45.15] | 58.97 | [54.85; | 62.98] | 20.65 | [14.12; | 29.18] | 79.35 | [70.82; | 85.88] | 56.02 | [49.01; | 62.80] | 43.98 | [37.20; | 50.99] |
| Alone / in a nursing home / hospital or other long-term care facility / other | 7.10 | [3.41; | 14.21] | 92.90 | [85.79; | 96.59] | 34.39 | [22.61; | 48.48] | 65.61 | [51.52; | 77.39] | 11.65 | [4.65; | 26.30] | 88.35 | [73.70; | 95.35] | 46.15 | [31.64; | 61.35] | 53.85 | [38.65; | 68.36] |
| Highest education level<sup>d</sup> |  |  |  |  |  |  |  |  |  |  |  |  |  |  |  |  |  |  |  |  |  |  |  |  |
| Did not attend school / primary level / not applicable / lower secondary level I / other | 8.89 | [3.92; | 18.92] | 91.11 | [81.08; | 96.08] | 43.90 | [29.10; | 59.87] | 56.10 | [40.13; | 70.90] | 26.09 | [13.97; | 43.41] | 73.91 | [56.59; | 86.03] | 62.22 | [45.11; | 76.75] | 37.78 | [23.25; | 54.89] |
| Upper secondary level II | 4.85 | [2.62; | 8.81] | 95.15 | [91.19; | 97.38] | 33.73 | [23.13; | 46.26] | 66.27 | [53.74; | 76.87] | 10.65 | [5.96; | 18.32] | 89.35 | [81.68; | 94.04] | 43.66 | [30.71; | 57.53] | 56.34 | [42.47; | 69.29] |
| Tertiary level | 6.64 | [2.76; | 15.11] | 93.36 | [84.89; | 97.24] | 37.44 | [30.11; | 45.40] | 62.56 | [54.60; | 69.89] | 19.70 | [9.98; | 35.18] | 80.30 | [64.82; | 90.02] | 53.10 | [40.97; | 64.87] | 46.90 | [35.13; | 59.03] |
| Medical specialty |  |  |  |  |  |  |  |  |  |  |  |  |  |  |  |  |  |  |  |  |  |  |  |  |
| Geriatrics | 1.67 | [1.67; | 1.67] | 98.33 | [98.33; | 98.33] | 5.08 | [5.08; | 5.08] | 94.92 | [94.92; | 94.92] | 1.79 | [1.79; | 1.79] | 98.21 | [98.21; | 98.21] | 8.33 | [8.33; | 8.33] | 91.67 | [91.67; | 91.67] |
| Gynecology | 12.38 | [9.69; | 15.68] | 87.62 | [84.32; | 90.31] | 46.74 | [38.68; | 54.97] | 53.26 | [45.03; | 61.32] | 34.69 | [27.24; | 42.98] | 65.31 | [57.02; | 72.76] | 69.52 | [67.45; | 71.52] | 30.48 | [28.48; | 32.55] |
| Internal medicine | 6.60 | [3.72; | 11.47] | 93.40 | [88.53; | 96.28] | 41.08 | [36.88; | 45.41] | 58.92 | [54.59; | 63.12] | 15.43 | [10.36; | 22.35] | 84.57 | [77.65; | 89.64] | 53.46 | [50.45; | 56.45] | 46.54 | [43.55; | 49.55] |
| Rehabilitation | 0.00 |  |  | 100.00 |  |  | 32.84 | [32.84; | 32.84] | 67.16 | [67.16; | 67.16] | 17.78 | [17.78; | 17.78] | 82.22 | [82.22; | 82.22] | 44.78 | [44.78; | 44.78] | 55.22 | [55.22; | 55.22] |
| Rheumatology | 2.56 | [2.56; | 2.56] | 97.44 | [97.44; | 97.44] | 36.84 | [36.84; | 36.84] | 63.16 | [63.16; | 63.16] | 12.50 | [12.50; | 12.50] | 87.50 | [87.50; | 87.50] | 46.15 | [46.15; | 46.15] | 53.85 | [53.85; | 53.85] |
| Missings stage 0: <sup>a</sup>173 missing values; <sup>b</sup>6 missing values; <sup>c</sup>88 missing values; <sup>d</sup>5 missing values; <sup>e</sup>135 missing values.<br>Missings stage 1: <sup>a</sup>160 missing values; <sup>b</sup>6 missing values; <sup>c</sup>84 missing values; <sup>d</sup>5 missing values; <sup>e</sup>126 missing values.<br>Missings stage 2: <sup>a</sup>99 missing values; <sup>b</sup>6 missing values; <sup>c</sup>62 missing values; <sup>d</sup>2 missing values; <sup>e</sup>89 missing values.<br>Missings stage 3: <sup>a</sup>88 missing values; <sup>b</sup>5 missing values; <sup>c</sup>56 missing values; <sup>d</sup>2 missing values; <sup>e</sup>76 missing values.<br>* Patients with psychosocial distress as indicated by scoring above cut-off at baseline in at least one of the three indicators: GAD-7, PHQ-8 and SSD-12.<br>Abbreviations: CHF, Swiss Francs; CI, 95% confidence interval; GAD-7, Generalized Anxiety Disorder (questionnaire with 7 items); ITT, intention to treat; LB, lower bound; PHQ-8, Depressive symptom scale with 8 items from the Patient Health Questionnaire; PPC, psychosomatic-psychiatric consultation; SSD-12, Somatic Symptom Disorder (questionnaire with 12 items); UB, upper bound. |  |  |  |  |  |  |  |  |  |  |  |  |  |  |  |  |  |  |  |  |  |  |  |  |

A planned survey design analysis was infeasible due to small cluster sizes in certain transition stages, which prevented reliable estimation of survey-adjusted variances.

Consequently, non-survey logistic regression models employing Huber–White robust standard errors were used to ensure model convergence and to obtain stable, comparable results across stages.

Multicollinearity was assessed through Variance Inflation Factors (VIFs), with all VIFs below 5, indicating no significant multicollinearity concerns. All statistics were generated using Stata/ SE (version 16 or later; StataCorp LP, College Station, TX, USA).

## 3. Results

A flowchart of study subjects is provided as supplementary, online only material 1. Analyses included 589 medical inpatients screening positive for distress (≥1 indicator above cutoff) and for whom, therefore, a PPC was considered. Table 1 summarizes the sociodemographic characteristics of these participants. Information on nationalities of the study participants is provided in more detail as supplementary, online only material 2.

Table 2 shows the detailed frequencies of transitioning across the four PPC service-use stages in the total sample and in the sample stratified according to predictor categories.

Overall, of the 589 patients who were *considered* for PPC, 93.9% were ultimately offered a PPC, respectively, a counseling session. Among the 553 patients who were offered a PPC, 63.1% accepted it. And of the 348 who accepted, 84.0% proceeded to at least one PPC session. Considering the entire sample of 589 from first consideration to final receipt of a consultation, approximately half of the distressed inpatients (50%) ultimately received a PPC, while the other half did not progress through to a consultation.

Results of the adjusted logistic regression models are presented in Table 3, with statistically significant predictions depicted in Figure 1. Nationality emerged as key predictor in several transitions: Patients without Swiss citizenship had significantly higher odds of moving from *Stage 1 (PPC offered) to Stage 2 (PPC accepted)* (OR = 1.82, 95% CI [1.10, 3.00]) as compared to Swiss citizens. In the overall transition from *Stage 0 (PPC considered) to Stage 3 (PPC received)*, having no Swiss citizenship predicted significantly higher odds of ultimately receiving a PPC (OR = 1.62 [1.01, 2.62]). The medical specialty of the ward was also strongly associated with the likelihood of progressing through multiple stages. Compared to patients in Internal Medicine (reference category), patients admitted in geriatrics showed markedly increased odds of *accepting* a PPC once it was offered (Stage 1 to Stage 2: OR = 14.77 [4.32, 50.41]) and of *receiving* it once accepted (Stage 2 to Stage 3: OR = 14.47 [1.80, 115.79]). They also had a higher likelihood of progressing from initial consideration (Stage 0) to receiving the PPC (Stage 3) (OR = 16.43 [5.98, 45.10]). By contrast, inpatients at gynecology wards had lower odds of transitioning from initial consideration (Stage 0) to having a PPC offered (to Stage 1) (OR = 0.21 [0.06, 0.63]) and from initial consideration (Stage 0) to receiving the PPC (Stage 3) (OR = 0.52 [0.28, 0.96]). For rheumatology wards, no consistent associations reached statistical significance. Small sample sizes and missing data limited the precision of certain ward specialty comparisons. Age, household income, marital status, living arrangement, highest educational level, and employment situation did not show consistent or significant associations with the transitions between PPC service-use stages. Although a few comparisons exhibited numerical increases or decreases in ORs, confidence intervals typically crossed unity, indicating no statistically robust relationship. The same was true for potential linear trends.

**Figure 1.**
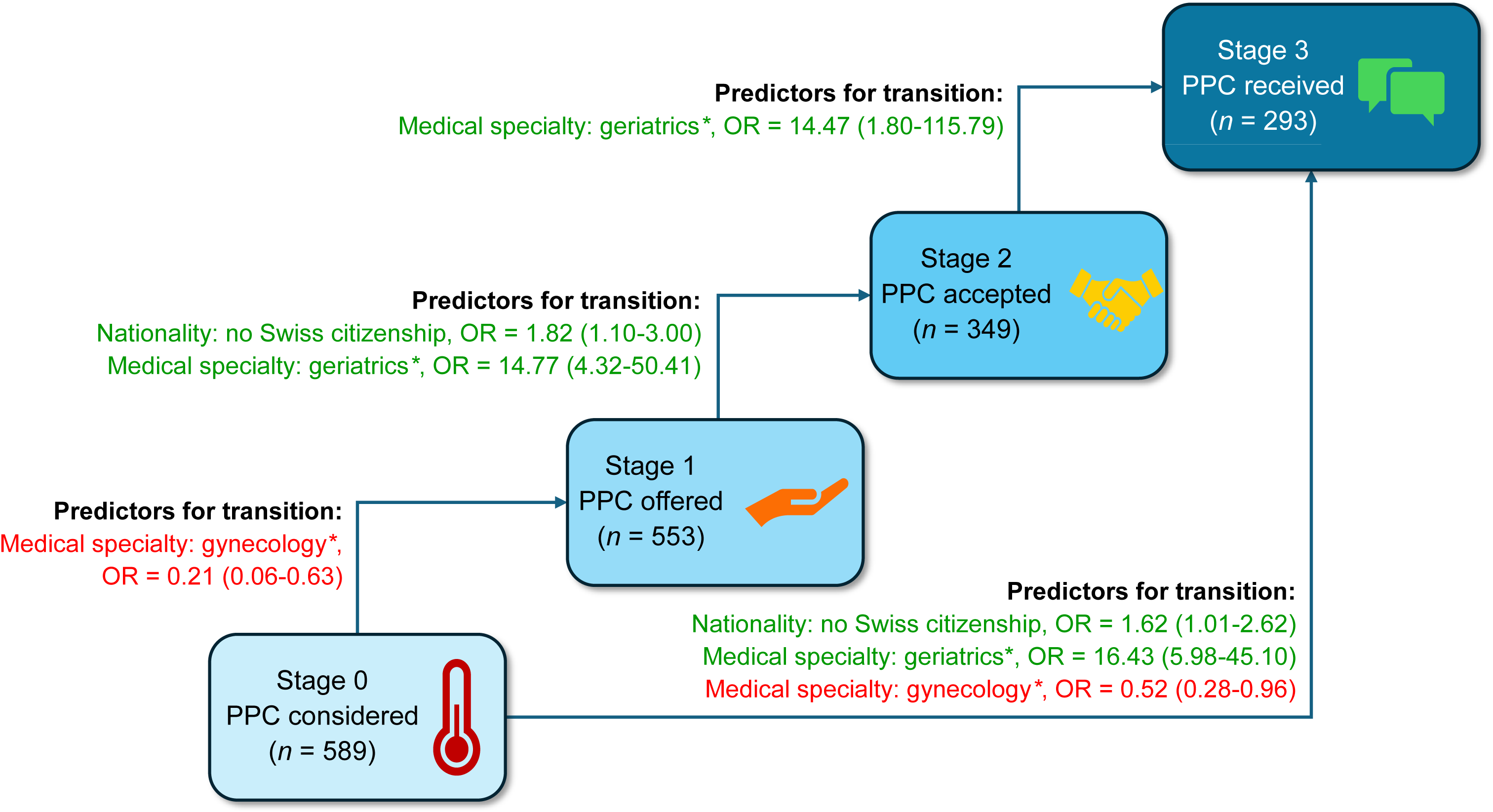
Patient journey from identified mental distress to receiving a PPC; and predictors for transitioning across the different stages. *\*versus internal medicine* *Notes: Only factors with statistically significant predictions are depicted (OR and 95%CI); For further details, see Table 3. Abbreviations: CI, confidence interval; OR, odds ratio; PPC, psychosomatic-psychiatric consultation*.

**Table 3.**
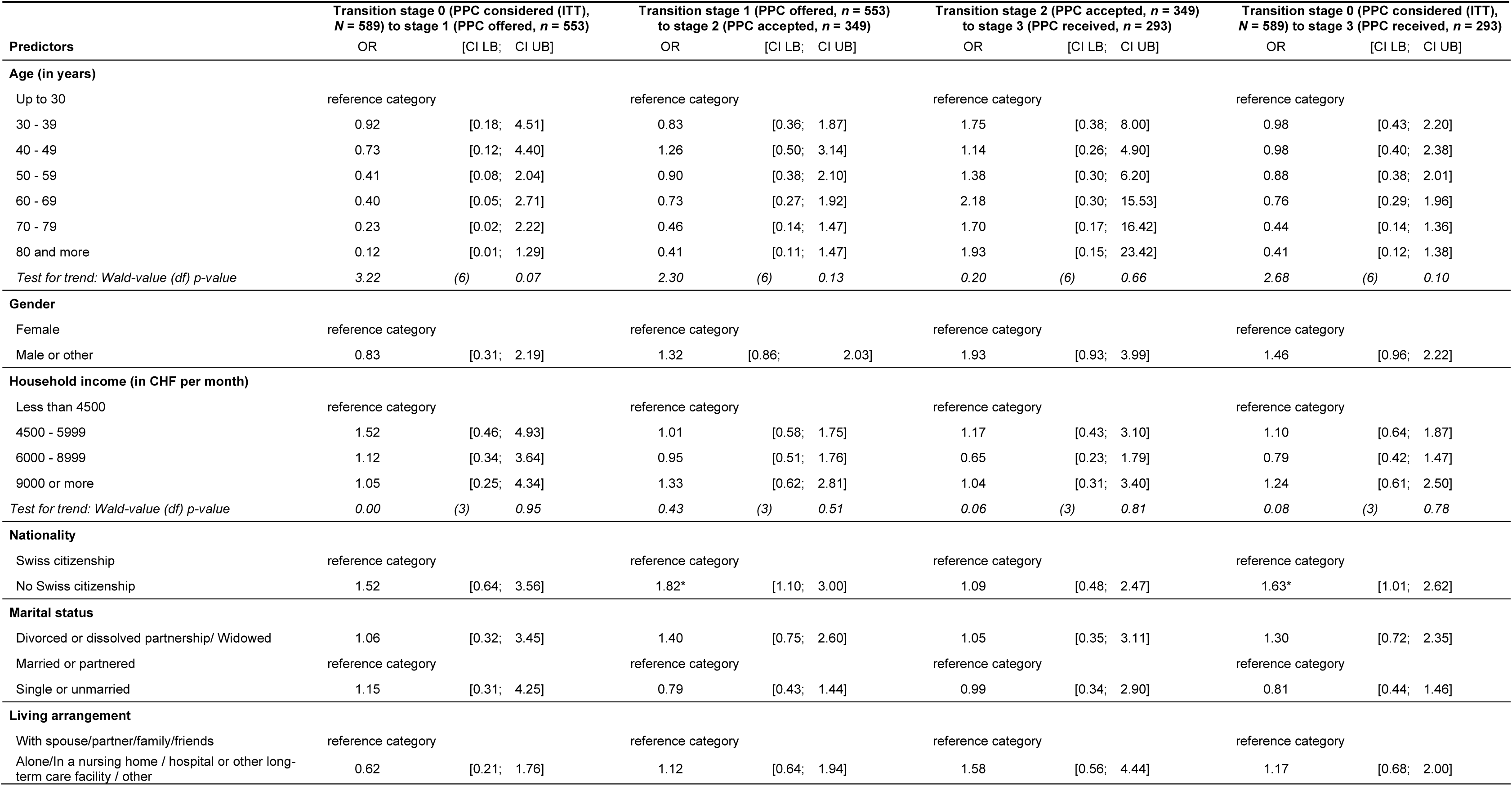
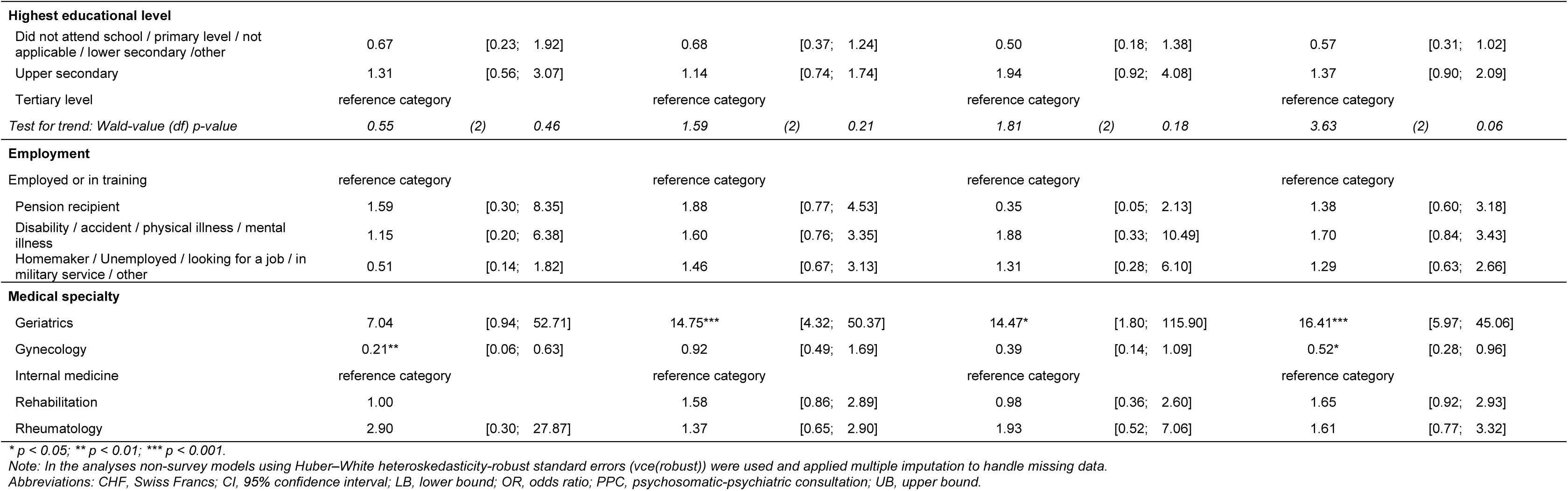
Results of logistic regression analyses with transitioning across the different PPC stages predicted by sociodemographic characteristics and medical specialty.

## 4. Discussion

### 4.1 Summary of main results

To the best of our knowledge, this is the first study to examine the uptake of psychosomatic-psychiatric consultations (PPCs) by medically hospitalized inpatients, specifically focusing on sociodemographic and ward-level predictors of progression through key stages of care. Nearly two thirds of inpatients offered a PPC accepted it. Only about half of mentally distressed inpatients ultimately received a PPC, highlighting limited access to mental healthcare during hospitalization. Two main predictors of PPC uptake emerged: Non-Swiss nationality was associated with a higher likelihood of acceptance and receipt. Ward medical specialty substantially shaped access: Patients in geriatric wards had markedly higher odds of progressing through all stages of care, while those in gynecology wards were significantly less likely to be offered or to receive a PPC.

### 4.2 Service uptake of PPCs in medically hospitalized inpatients

Our findings highlight that almost two-thirds of patients offered a PPC accepted it. This is notably higher than uptake rates reported in other outpatient settings. For instance, Frey Nascimento and colleagues [14] found that only 23% of distressed cancer outpatients attended a psycho-oncological service within four months after their first oncology consultation. In that study, uptake was strongly predicted by whether the oncologist explicitly recommended the service, whereas simply discussing distress or providing information had no significant effect. A multi-site Austrian study of 728 general hospital inpatients reported that 34.2% were cases with mental disorders; among these, 51.8% were judged to require either psychiatric consultation or inpatient referral, yet only 33.3% of those with an indicated need actually received a consultation [15]. The comparatively high acceptance (63.1%) and receipt rates (50%) in our sample may thus reflect not only the structured implementation of stepped-care pathways in the SomPsyNet model, but also the effectiveness of proactive, direct offers of PPC within the medical hospital inpatient context, which reach patients well due to their very low-threshold nature. This approach aligns with proactive integrated consultation-liaison psychiatry, which embeds early screening and co-located care into routine ward practice rather than relying on reactive referrals [16].

### 4.3 Missed transitions: From distress screening to received PPC

Nevertheless, in our study about one-sixth of those who accepted did not ultimately receive the PPC. The fact that about half of distressed inpatients never progressed to receive a PPC emphasizes the need to identify and address whatever is impeding progress at each step of care. In our data, the only significant factors associated with dropouts were patient nationality and ward medical specialty, suggesting that efforts to improve uptake may concentrate on these areas.

### 4.4 Non-Swiss citizenship as predictor of PPC uptake

Patients without Swiss citizenship were more likely to accept and ultimately receive a PPC than Swiss citizens. This finding is noteworthy, given that the study included only German-speaking participants, thereby excluding individuals with insufficient German proficiency. The non-Swiss subgroup predominantly comprised German-speaking migrants from Germany and other Western European countries, indicating high linguistic and cultural proximity to the Swiss healthcare system. The observed association is therefore unlikely to be driven by language-related barriers or by vulnerabilities typically associated with labor migration.

Population-based evidence from Switzerland does not indicate higher physician consultation rates among non-Swiss residents with comparable linguistic and cultural backgrounds. On the contrary, non-migrants are generally more likely to visit a physician than first-generation or culturally different immigrants, while consultation rates among culturally similar immigrants are comparable – but not higher – than those of Swiss nationals [17]. Against this background, our findings likely reflect mechanisms specific to the proactive, low-threshold consultation-liaison context of medically hospitalized inpatient care. Higher psychosocial distress among migrant populations [17] and structured care arrangements designed to reduce access barriers [18] may facilitate both the offering and uptake of PPCs once distress is identified. Given mixed evidence on mental health service utilization among non-citizen groups [19], our results suggest that proactive consultation-liaison models embedded in routine inpatient care can mitigate barriers that might otherwise limit psychosomatic or psychiatric service uptake among migrant or minority patients.

### 4.5 Medical specialty of the ward as determinant of PPC uptake

In line with prior studies on consultation-liaison psychiatry [6], our data show that specialty of wards can substantially shape the likelihood of a patient receiving mental healthcare. Qualitative findings from the larger context of the SomPsyNet study also suggest that organizational culture and specific ward routines are relevant when addressing mental-somatic comorbidity [18]. Patients in geriatric wards had a markedly higher likelihood of transitioning from “PPC offered” to “PPC accepted” and from “PPC accepted” to “PPC received.” Geriatric care often includes multidisciplinary teams skilled at recognizing the interplay of somatic and mental problems, thereby facilitating patient willingness and readiness to engage in psychosomatic or psychiatric care. Moreover, geriatric patients often exhibit greater clinical complexity, such as multiple comorbid conditions, which has previously been shown to be associated with increased utilization of mental health services within SCCMs [19]. Furthermore, older adults are more likely to adhere to physicians’ and health carer recommendations [20,21].

Conversely, our findings suggest that patients in gynecology wards were significantly less likely to be offered or ultimately receive a PPC. This finding is in line with previous studies reporting notably low referral rates for psychiatric consultations among obstetric and gynecology inpatients [22]. This pattern may be attributable to practical factors, such as shorter hospital stays, a younger patient population, and differing staff practices that reduce opportunities for referral.

These results align with previous observations that organizational culture and workflow integration within certain specialties are decisive in bridging patients to mental health interventions [23]. Notably, we found no indicators of effects of patient age or sex on PPC uptake, in contrast to some prior studies [24]. This discrepancy could be due to our exclusion of certain populations (e.g., oncology patients) or the overriding influence of the ward context. In summary, beyond ward specialty and nationality, we did not identify other significant predictors of whether an inpatient received psychosomatic-psychiatric care.

### 4.6 Strengths and limitations

A key strength of our study is its relatively large and diverse sample, recruited from multiple wards in a large, heterogeneous cohort (N=589) from three medical hospitals. This diversity supports the representativeness of our findings across a wide spectrum of medically hospitalized inpatients. Additionally, the stepped-wedge cluster randomized design provides a pragmatic approach to evaluating how patients move through different levels of stepped and collaborative care in real-world hospital settings.

However, several limitations must also be noted. First, the SomPsyNet study did not include the full range of medical specialties. For example, we did not include surgical wards (of note, some patients, e.g., from gynecology wards, may still have undergone surgery) or wards with patients primarily treated for oncological conditions, as for these patients, specific psychosocial support services are already available. Second, the inclusion of gynecology may have introduced some degree of gender imbalance, although we adjusted for gender in all analyses. Third, the study focused solely on German-speaking participants, potentially excluding those who lacked sufficient German proficiency. Therefore, our finding of higher PPC uptake among non-Swiss nationals pertains only to individuals without major language barriers; it may not apply to patients who face significant language or communication challenges. Finally, we cannot rule out a selection bias: patients who felt better or worse than average might have been more inclined (or had more opportunity) to participate.

### 4.7 Generalizability across health systems and clinical settings

With regard to generalizability, it shall be noted that our study was conducted in urban medical hospitals within a country with a highly developed healthcare system. The structure, resources, and organizational culture in Swiss medical hospitals differ from settings with fewer resources or different healthcare models. Consequently, caution should be exercised when transferring these findings to regions where psychosomatic or psychiatric care is less integrated into routine hospital practice. Moreover, although we included wards focusing on a range of medical specialties, our omission of surgical units and the unique patient flow patterns in gynecology may limit extrapolation to other hospital contexts.

### 4.8 Implications for clinical practice, policy, and future research

The results of our study allow deriving several implications. From a clinical perspective, our findings highlight the importance of systematically integrating mental health care – delivered through PPCs – into medical wards. Such integration inherently includes routine, standardized screening for psychosocial distress to enable early identification of patients in need of support. This approach aligns with proactive integrated consultation-liaison psychiatry, which embeds early screening and co-located mental health care into routine ward practice rather than relying on reactive, ad hoc referrals [16]. In practice, this is best achieved through well-established liaison models.

Building on this integrated, proactive framework, targeted strategies should focus on units with consistently lower uptake by addressing unit-specific logistical barriers, providing staff training, and embedding mental health professionals in day-to-day ward routines to facilitate referrals. Moreover, once a patient accepts a PPC, timely delivery during the hospital stay appears crucial. Streamlining the referral-to-consultation process – for example, by scheduling a PPC at the earliest opportunity, as well as ensuring adequate staffing of the consultation-liaison psychosomatic/psychiatric team – may help prevent patients from missing needed support due to discharge or procedural delays. Additionally, as the largest drop-off occurred when patients declined the offered PPC, efforts to de-stigmatize mental health services and to educate patients about the benefits of PPC may be prioritized to improve acceptance.

Policymakers could consider incentivizing or mandating close collaboration between specialist wards and mental health services to ensure consistent and transparent referral pathways. Policies that support the implementation of proactive consultation-liaison models – including routine screening, co-located mental health professionals, and adequate staffing – may help reduce unmet psychosocial needs and improve equitable access to mental health care in medical hospitals.

Scientifically, our study underscores that future research should account for institutional factors – including ward culture, staff perceptions of psychosocial distress, and organizational workflows – when evaluating the uptake of psychosomatic-psychiatric services. In addition, future analyses should examine the health economic implications of integrated and proactive consultation-liaison approaches. Linked insurance claims data – partly already available and partly forthcoming within our study – allow assessment of short-term and longer-term patterns of health care use and related outcomes. Future research in geographically diverse and resource-varied environments is essential to determine whether our findings hold true or differ systematically.

### 4.9 Conclusions

In conclusion, nearly two-thirds of medically hospitalized inpatients accepted a PPC when it was offered. About half ultimately received a consultation, indicating substantial attrition along the stepped and collaborative care pathway. Nationality and ward specialty were key determinants of progression, with higher uptake in Geriatric wards and lower uptake in Gynecology. The higher PPC uptake among non-Swiss patients should be interpreted in the context of proactive, low-threshold inpatient consultation-liaison care rather than as a general pattern of healthcare utilization. In line with national strategies addressing mental health in the context of non-communicable diseases [10] and ongoing efforts to integrate somatic and mental healthcare [27], these findings highlight the need for ward-oriented, structurally embedded consultation-liaison models. Systematic screening, close interprofessional coordination, and co-located mental health services may promote timely, equitable, and effective mental health services for medically hospitalized inpatients, including migrant and minority patients who are otherwise less likely to access such care.

## Supporting information

Flow chart and suppl. table 1

## Data availability

The datasets being held by the project are not readily available due to their sensitive nature. In the case of inquiries by third parties that wish to reuse the project’s data after an embargo period, the following procedure is planned. Researchers interested in the data may submit a project synopsis addressed to the publications committee of the project and will have to obtain authorization from the responsible ethics committee as ordained in the Ordinance of 20 September 2013 on Human Research with the exception of Clinical Trials [25,26] (Human Research Ordinance, HRO). The publication committee will review the project synopsis and will answer the formal requests of applicants. Only upon collection of all important consents and upon approval of the responsible ethics committee(s), the requested data will be transferred to the applicants. Third parties must confirm and provide evidence to comply with all relevant Swiss and cantonal laws and regulations (especially regarding data protection and Human Research), as well as all obligations and regulations set out in the documents and contracts related to the project. Fees may apply to cover expenses related to data reuse. Requests to access the datasets should be directed to the corresponding author. Any future changes to the data sharing plan will be noted in Data Availability Statements and updated in the registry record.

## Author contributions including CRediT statement

The coordinating center at UHB oversaw all activities across all sites. The SomPsyNet steering committee included the project lead and operations manager of the Department of Health, Canton Basel-Stadt/ Division of Prevention, the study sponsor, the principal investigators, the operations manager at the coordinating center, and a representative of the St. Claraspital Medical Clinic (CLARA), which participated in the SomPsyNet project but not in the SomPsyNet evaluation study. The steering committee was responsible for all major study decisions. The endpoint was discussed among the investigators of the steering committee, with additional input from members of the co-author group. Some of these (KW, GF, MS) also served on the external evaluation board, which evaluated SomPsyNet.

Health Promotion Switzerland (GFCH), the main funding body of SomPsyNet, provided the project funding.

An advisory board provided guidance and feedback related to the project and the study. A patient advisory group consisting of several patient representatives worked together to oversee and give feedback on study materials and protocol. The SomPsyNet consortium met at least on a yearly basis, discussing the development of the study, and giving critical feedback on the study conduction and necessary adaptations.

CK, RS, AF, and GM first identified the question leading to the formation of this research. GM, AF, CK, AS, IB, MB, AD, STs, KW, GF, MS, and RS contributed to the development of the main trial protocol. RS contributed significantly also as the sponsor-investigator of this study. GM served as lead principal investigator, and AF, MB, RS, STr, and STs served as principal investigators at the different study sites, contributing significantly to data collection and protocol adherence. IB, and AS acted as study operative leads, overseeing and managing the execution of the study. GM drafted the first version of the manuscript. SB, and NC conducted statistical analyses. IB, LE, NC, and DB were in charge and conducted data management tasks. All authors made significant revisions to the manuscript for important intellectual content, and all authors reviewed and approved the final version of the manuscript for submission, reflecting their agreement with the work in its current form and their acceptance of accountability for all aspects of the work. This manuscript was written following the STROBE reporting guidelines.

## Declaration of Competing Interest & Funding Independent of the Project

GM & RS received funding in the context of a Horizon Europe project from the Swiss State Secretariat for Education, Research and lnnovation (SERI) under contract number 22.00094. Further, GM & RS received funding from Wings Health Inc. in the context of a proof-of-concept study. GM received funding from the Swiss Heart Foundation under project no. FF21101, from the Research Foundation of the International Psychoanalytic University (IPU) Berlin under projects no. 5087 and 5217, from the Swiss National Science Foundation (SNSF) under project no. 100014_135328, from the German Federal Ministry of Education and Research under budget item 68606, and from the Hasler Foundation under project no. 23004. GM is co-founder and holds stock in Therayou AG, which is active in the field of digital and blended mental healthcare. GM receives royalties from publishing companies as author, including a book published by Springer, and an honorarium from Lundbeck for speaking at a symposium. Furthermore, GM is compensated for providing psychotherapy to patients, acting as a supervisor, serving as a self-experience facilitator (‘*Selbsterfahrungsleiter*’), and for postgraduate training of psychotherapists and supervisors. RS received a speaker honorarium from Novartis. RS is chairman of the Basel Institute for Psychosomatic Medicine (BIPM) and founder and managing director of the Psychosomatic and Psychosocial Services GmbH, that develops and implements psychosomatic and psychosocial training and continuing education programs. He is a member of the board of trustees of the Foundation Psychosomatic and Social Medicine (Ascona Foundation).

The authors declare no other potential conflict of interest. The research activities were fully independent, and there were no intellectual or financial proprietary claims.

## Declaration of generative AI and AI-assisted technologies

We used artificial intelligence (AI)-based tools, including ChatGPT and OpenEvidence to support manuscript preparation. Further, we used publicly available search technologies, which we recognize likely utilize AI capabilities. We confirm that the contributions of AI were strictly in an assistive capacity. AI was not involved in conceptual tasks. Human oversight was continuously employed to ensure the accuracy of content and address any ethical concerns.

## MEMBERS OF THE SOMPSYNET CONSORTIUM

Bachmann, M., Department of Psychosomatic Medicine and Psychotherapy, Klinik Barmelweid AG, Barmelweid, Switzerland; Baenteli, I., Department of Psychosomatic Medicine, University Hospital and University of Basel, Basel, Switzerland; Bahmane, S., Department of Psychosomatic Medicine, University Hospital and University of Basel, Basel, Switzerland; Bales, G., University Department of Geriatric Medicine FELIX PLATTER, Basel, Switzerland; Bally, K., Centre for Primary Health Care, University of Basel, Switzerland; Bassetti, S., Division of Internal Medicine, University Hospital and University of Basel, Basel, Switzerland, Department of Clinical Research, University Hospital and University of Basel, Basel, Switzerland; Baumgartner, R., Social Insurance Institution Basel-Landschaft, Binningen, Switzerland; Beck, J., Clinic Sonnenhalde AG, Riehen, Switzerland; Bosman, S., Department of Psychosomatics and Psychiatry, Bethesda Hospital, Basel, Switzerland.; Buechel, D., University of Basel c/o University Hospital of Basel, Department of Clinical Research, Basel, Basel-Stadt, Schweiz; Dörner, A., St. Claraspital, Medical clinic, Basel, Switzerland; Ebner, L., Department of Psychosomatic Medicine, University Hospital and University of Basel, Basel, Switzerland; Erb, J., Department of Psychosomatic Medicine, University Hospital and University of Basel, Basel, Switzerland; Fink, G., Swiss Tropical and Public Health Institute, Basel, Switzerland; University of Basel, Basel, Switzerland; Frick, A., Department of Psychosomatic Medicine, University Hospital and University of Basel, Basel, Switzerland; Fuchs, S., Department of Health Canton Basel-Stadt, Medical Services, Basel, Switzerland; Grossmann, F., Department of Medicine, Division of Nursing, University Hospital Basel, Basel, Switzerland; Hermann, A., Direktion Pflege/MTT, University Hospital and University of Basel, Basel, Switzerland; Hotopf, M., Department of Psychological Medicine, Institute of Psychiatry, Psychology and Neuroscience, King’s College London, London, United Kingdom; Huber, C., University Psychiatric Clinics (UPK), University of Basel, Basel, Switzerland; Isler-Christ, L., Sevogel-Apotheke, Basel, Switzerland, Baselstädtischer Apotheker-Verband, Basel, Switzerland; Karpf, C., Department of Health Canton Basel-Stadt, Division of Prevention, Basel, Switzerland; Katapodi, MC., Department of Clinical Research, University of Basel, Basel, Switzerland, University of Michigan School of Nursing, Ann Arbor, MI USA; Keller, RC., Swiss Heart Foundation, Bern, Switzerland; Klimmeck, S., University Hospital Basel, Basel, Switzerland; Lang, UE., University Psychiatric Clinics (UPK), Department of Psychiatry and Psychotherapy, Basel, Switzerland; Mazander, S., IV-Stelle Basel-Stadt, Basel, Switzerland; Meinlschmidt, G., Department of Digital and Blended Psychosomatics and Psychotherapy, Psychosomatic Medicine, University Hospital and University of Basel, Basel, Switzerland; Department of Psychosomatic Medicine, University Hospital and University of Basel, Basel, Switzerland; Trier University, Department of Clinical Psychology and Psychotherapy – Methods and Approaches, Trier, Germany; Schaefert, R., Department of Psychosomatic Medicine, University Hospital and University of Basel, Basel, Switzerland; Schirmer, F., Vereinigung der psychosomatisch tätigen Aerztinnen und Aerzte der Region Basel, Basel, Switzerland; Schur, N., Institute of Pharmaceutical Medicine (ECPM), University of Basel, Basel, Switzerland; Schwenkglenks, M., Institute of Pharmaceutical Medicine (ECPM), University of Basel, Basel, Switzerland, Health Economics Facility, Department of Public Health, University of Basel, Basel, Switzerland; Schwob, P., Psychotherapists Association of Basel VPB, Basel, Switzerland; Seelmann, SF., Department of Internal Medicine, University Hospital Basel, Basel, Switzerland; Stiefel, F., Liaisonpsychiatrischer Dienst, University Hospital Lausanne, Lausanne, Switzerland; Studer, A., Department of Health Canton Basel-Stadt, Division of Prevention, Basel, Switzerland; Tegethoff, M., Institute of Psychology, RWTH Aachen University, Aachen, Germany; Trost, S., Department of Geriatric Psychiatry, Universitäre Altersmedizin FELIX PLATTER, Basel, Switzerland; Tschudin, S., Department of Obstetrics and Gynecology, University Hospital and University of Basel, Switzerland; Urech, C., Gyn. Social Medicine and Psychosomatics, University Hospital and University of Basel, Basel, Switzerland; Weyermann, D., Patientenstelle Basel, Basel, Switzerland; Wyss, K., Swiss Tropical and Public Health Institute, Basel, Switzerland; University of Basel, Basel, Switzerland; Zwahlen, D., Department of Psychosomatic Medicine, University Hospital and University of Basel, Basel, Switzerland; von Allmen, T., Department of Health Canton Basel-Stadt, Health Care, Basel, Switzerland

## Acknowledgements

The authors thank all patients who participated in the study and generously shared their experiences, as well as all members of the consortium involved in providing support in patient recruitment at the various study sites. Further, we are grateful to all master’s students involved for their assistance in recruitment and overall study implementation. Special thanks go to the patient participation committee for their valuable input in grant preparation, study design, refinement of consent materials, and strategies for enrolment and dissemination. We thank the physicians, nursing and other staff from all participating units at each study site for their support throughout the study. We also acknowledge the contributions of the involved ICT specialists, whose technical support was essential. We would like to thank the Canton of Basel-Stadt/ Department of Health/ Division of Prevention, and all project partners for their excellent collaboration and contribution. We are very grateful to Health Promotion Switzerland for their continued funding and support. We thank our advisory board for their valuable advice and the external evaluation team of the SomPsyNet Study for their thoughtful accompaniment. Finally, our sincere thanks go to all members of the SomPsyNet consortium for their expertise and constructive collaboration during all phases of the project.

## STROBE Statement—checklist of items that should be included in reports of observational studies

XXX ⇒ the page numbers will be updated, once the final edited article is ready

|  | Item No. | Recommendation | Page No. | Relevant text from manuscript |
| --- | --- | --- | --- | --- |
| **Title and abstract** | 1 | (a) Indicate the study's design with a commonly used term in the title or the abstract | 1-2 | see abstract |
|  |  | (b) Provide in the abstract an informative and balanced summary of what was done and what was found | 1-2 | see abstract |
| **Introduction** |  |  |  |  |
| Background/rationale | 2 | Explain the scientific background and rationale for the investigation being reported | 4-6 | see introduction section |
| Objectives | 3 | State specific objectives, including any prespecified hypotheses | 5-6 | see introduction section |
| **Methods** |  |  |  |  |
| Study design | 4 | Present key elements of study design early in the paper | 6 | see methods section, paragraph "Study setting and design" |
| Setting | 5 | Describe the setting, locations, and relevant dates, including periods of recruitment, exposure, follow-up, and data collection | 7 | see methods section, paragraphs "Study setting and design" and "Inclusion and exclusion criteria" |
| Participants | 6 | *Cross-sectional study*—Give the eligibility criteria, and the sources and methods of selection of participants | 7-8 | see methods section, paragraph "Inclusion and exclusion criteria" |
| Variables | 7 | Clearly define all outcomes, exposures, predictors, potential confounders, and effect modifiers. Give diagnostic criteria, if applicable | 8 | see methods section, paragraph "study variables" |
| Data sources/measurement | 8 | For each variable of interest, give sources of data and details of methods of assessment (measurement). Describe comparability of assessment methods if there is more than one group | 8-9 | see methods section, paragraph "study variables" |
| Bias | 9 | Describe any efforts to address potential sources of bias | 8-9 | see methods section, paragraph "study variables" and "data collection" |
| Study size | 10 | Explain how the study size was arrived at | 8-9 | see methods section, paragraph “study variables” and “data collection” |
| Quantitative variables | 11 | Explain how quantitative variables were handled in the analyses. If applicable, describe which groupings were chosen and why | 9-11 | see methods section, paragraph “statistical methods” |
| Statistical methods | 12 | (a) Describe all statistical methods, including those used to control for confounding | 9-11 | see methods section, paragraph “Statistical methods” |
|  |  | (b) Describe any methods used to examine subgroups and interactions |  |  |
|  |  | (c) Explain how missing data were addressed | 10 | see methods section “Statistical methods” |
|  |  | (d) *Cross-sectional study*—If applicable, describe analytical methods taking account of sampling strategy | not applicable |  |
|  |  | (e) Describe any sensitivity analyses | not applicable |  |
| **Results** |  |  |  |  |
| Participants | 13 | (a) Report numbers of individuals at each stage of study—e.g. numbers potentially eligible, examined for eligibility, confirmed eligible, included in the study, completing follow-up, and analyzed | 12 | see results section, supplementary, online only material 1 |
|  |  | (b) Give reasons for non-participation at each stage | 12 | see results section, supplementary, online only material 1 |
|  |  | (c) Consider use of a flow diagram | 12 | see results section, supplementary, online only material 1 |
| Descriptive data | 14 | (a) Give characteristics of study participants (e.g. demographic, clinical, social) and information on exposures and potential confounders | 12-19 | see results section, tables 1, 2, and 3 |
|  |  | (b) Indicate number of participants with missing data for each variable of interest | 12-19 | see results section, tables 1, 2, and 3 |
| Outcome data | 15 | *Cross-sectional study*—Report numbers of outcome events or summary measures | 19 | see results section, figure 1 |
| Main results | 16 | (a) Give unadjusted estimates and, if applicable, confounder-adjusted estimates and their precision (e.g. 95% confidence interval). Make clear which confounders were adjusted for and why they were included | 12-19 | see results section and tables 2 and 3 |
|  |  | (b) Report category boundaries when continuous variables were categorized | 12-15 | see results section, table 2 |
|  |  | (c) If relevant, consider translating estimates of relative risk into absolute risk for a meaningful time period | not relevant |  |
| Other analyses | 17 | Report other analyses done - e.g. analyses of subgroups and interactions, and sensitivity analyses | 12-15 | see results section |
| **Discussion** |  |  |  |  |
| Key results | 18 | Summarize key results with reference to study objectives | 20-23 | see discussion section, paragraph “summary of main results” |
| Limitations | 19 | Discuss limitations of the study, taking into account sources of potential bias or imprecision. Discuss both direction and magnitude of any potential bias | 24-25 | see discussion section, paragraph “Strengths and Limitations” |
| Interpretation | 20 | Give a cautious overall interpretation of results considering objectives, limitations, multiplicity of analyses, results from similar studies, and other relevant evidence | 20-25 | see discussion section, paragraph “Clinical, public health, and future research implications” |
| Generalizability | 21 | Discuss the generalizability (external validity) of the study results | 25 | see discussion section, paragraph “Generalizability” |
| **Other information** |  |  |  |  |
| Funding | 22 | Give the source of funding and the role of the funders for the present study and, if applicable, for the original study on which the present article is based | 10 (part 1) | see “Funding sources” and “Declaration of Competing Interest & Funding Independent of the Project” |

