## Supplementary material for "Patient- and Ward-Level Determinants of Psychosomatic-Psychiatric Consultations for Mentally Distressed Inpatients from Medical Hospitals: Findings from the SomPsyNet Stepped-Wedge-Trial": Flow chart and suppl. table 1

### SUPPLEMENTARY, ONLINE ONLY MATERIALS

#### Supplementary, online only material 1. Flow chart of study participants.

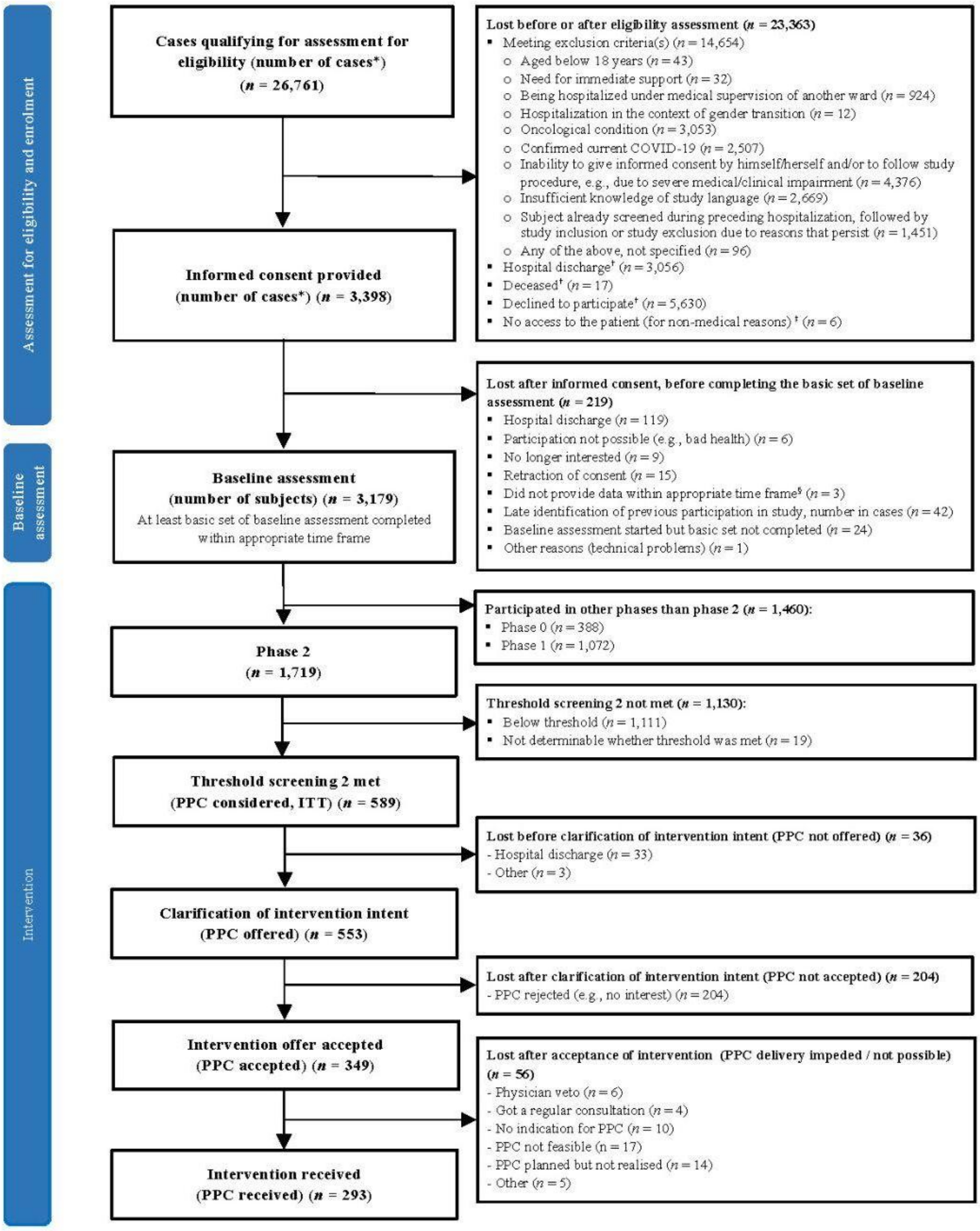


*Supplementary, online only material 1. Flow chart of study participants.*

*Due to multiple study centers, repeated recruitment and inclusion of the same patient could not always be prevented. Therefore, numbers are here shown in cases (i.e., the same subject could contribute to several cases).

†Not confirmed that we have no exclusion criteria.

‡Patients under police/security surveillance, who were not recruited for employee safety reasons.

§Completion of baseline assessment > 30 days after hospital discharge.

Abbreviations: COVID-19, Coronavirus disease 2019; ITT, intention to treat; PPC, psychosomatic-psychiatric consultation.

#### Supplementary, online only material 2. Information on nationalities of the study participants.

**Supplementary, online only material 2. Information on nationalities of the study participants.**

| Category (WHO region[a]) | Country/area | n[c,d] | % of total (N=588)[c,d] | % of non-Swiss (n=121)[c,d] |
|---|---|---|---|---|
| **European Region[b] (EUR, excluding Switzerland)** | | **112** | **19.0%** | **92.6%** |
| | **Western Europe** | **71** | **12.1%** | **58.7%** |
| | Germany | 58 | 9.9% | 47.9% |
| | Other countries | 13 | 2.2% | 10.7% |
| | **Southern Europe** | **27** | **4.6%** | **22.3%** |
| | Italy | 12 | 2.0% | 9.9% |
| | Other countries | 15 | 2.6% | 12.4% |
| | **Other European Regions[e]** | **14** | **2.4%** | **11.6%** |
| **Other Regions** (Region of the Americas, Western Pacific Region, SouthEast Asia Region, Eastern Mediterranean Region, African Region) | | **12** | **2.0%** | **9.8%** |

[a] *by World Health Organization (2024). Countries and areas: WHO regional groupings. Geneva: WHO. Available at: https://www.who.int/countries*

[b] *by United Nations, Department of Economic and Social Affairs, Statistics Division (2021). Standard Country or Area Codes for Statistical Use (M49). Series M, No. 49. New York: United Nations. Available at: https://unstats.un.org/unsd/methodology/m49/*

[c] *one non-swiss participant with missing information.*

[d] *3 non-swiss participants mentioned two nationalities, hence numbers may not sum up to 100%.*

[e] *Other European Regions here comprise Eastern Europe, Northern Europe and Western Asia; of note, Israel and Türkiye are WHO/Europe members but are Western Asia in the UN M49 geoscheme, so they are here assigned to Western Asia.*

*Abbreviations: WHO, World Health Organisation.*

*Note. In accordance with privacy-preserving practices, any nationality subgroup containing fewer than ten participants was suppressed. These subgroups were collapsed and reported as 'Other' entity.*
